# Critical Role of the Subways in the Initial Spread of SARS-CoV-2 in New York City

**DOI:** 10.1101/2021.07.03.21259973

**Authors:** Jeffrey E. Harris

## Abstract

We studied the possible role of the subways in the spread of SARS-CoV-2 in New York City during late February and March 2020. Data on cases and hospitalizations, along with phylogenetic analyses of viral isolates, demonstrate rapid community transmission throughout all five boroughs within days. The near collapse of subway ridership during the second week of March was followed within 1-2 weeks by the flattening of COVID-19 incidence curve. We observed persistently high entry into stations located along the subway line serving a principal hotspot of infection in Queens. We used smartphone tracking data to estimate the volume of subway visits originating from each zip code tabulation area (ZCTA). Across ZCTAs, the estimated volume of subway visits on March 16 was strongly predictive of subsequent COVID-19 incidence during April 1-8. In a spatial analysis, we distinguished between the conventional notion of geographic contiguity and a novel notion of contiguity along subway lines. We found that the March 16 subway-visit volume in subway-contiguous ZCTAs had an increasing effect on COVID-19 incidence during April 1-8 as we enlarged the radius of influence up to 5 connected subway stops. By contrast, the March 31 cumulative incidence of COVID-19 in geographically-contiguous ZCTAs had an increasing effect on subsequent COVID-19 incidence as we expanded the radius up to 3 connected ZCTAs. The combined evidence points to the initial citywide dissemination of SARS-CoV-2 via a subway-based network, followed by percolation of new infections within local hotspots.

## Introduction

An accurate, thorough understanding of the rapid, widespread propagation of SARS-CoV-2 infection during the early phase of the massive outbreak in New York City is crucial to the successful control of future pandemic threats.

To that end, we test three main hypotheses here. First, New York City’s extensive public transport system, particularly its subways, played a critical role in the widespread dissemination of SARS-CoV-2 infection throughout the city during the end of February and the beginning of March 2020. Second, the ensuing marked decline in subway use was an important vehicle by which the public’s growing perception of risk was translated into reduced community transmission of the virus. Third, those areas with an attenuated decline in subway use, we posit, subsequently became the loci for high-density clusters of viral infection in late March 2020.

The Metropolitan Transportation Authority (MTA), a network of subways, buses and commuter rail cars serving the NYC area, is larger than all other metropolitan transport systems in the United States combined. While nearly 85 percent of U.S. workers drive to their jobs, according to the MTA, 80 percent of rush-hour commuters to the city’s central business districts use transit.^1^ The MTA’s subway system is particularly unique, with a total of 1697.8 million turnstile entries during the calendar year 2019,^2^ compared to 157.2 million entries into the Washington DC metro,^3^ the next largest subway system in the country.

Our hypotheses are hardly novel. The role of transportation networks in the spread of SARS-CoV-2 has been supported by recent studies of the initial outbreak in Wuhan, China.^4-6^ One study of the NYC epidemic found an association between continued subway use among essential workers and a delayed flattening of the epidemic curve.^7^ Another study based in part on NYC subway ridership data found a link between mobility and COVID-19 risk.^8^ Yet another study found strong correlations between NYC subway turnstile entries and COVID-19 cases and deaths.^9^

What sets our study apart is its comprehensive, multidisciplinary approach. We rely on such diverse lines of evidence as phylogenetic analysis of early viral samples, public health data on confirmed COVID-19 cases, public transport data on turnstile entries, location-tracking data on the movements of smartphones, and census data on the prevalence of at-risk multi-generational households. Our spatial analysis of emerging case clusters distinguishes critically between the conventional notion of geographic contiguity and what we call *subway contiguity*.

## Materials and Methods

### Data on Confirmed COVID-19 Cases

The NYC health department’s open data archive^10^ was our source of data on: confirmed COVID-19 cases and hospitalizations by borough and date of diagnosis (*boroughs-case-hosp-death*, used to construct Figs. 1a, 1b and 2b below), aggregate, city-wide data on cases and hospitalizations by date of diagnosis (*case-hosp-death*, used in part to construct Fig. 2a); and cumulative cases by zip code tabulation area (ZCTA) (*tests-by-zcta*, used in part to construct Figs. 2c and 3d). Incidence per 10,000 population was based on population counts described below.

**Fig. 1:**
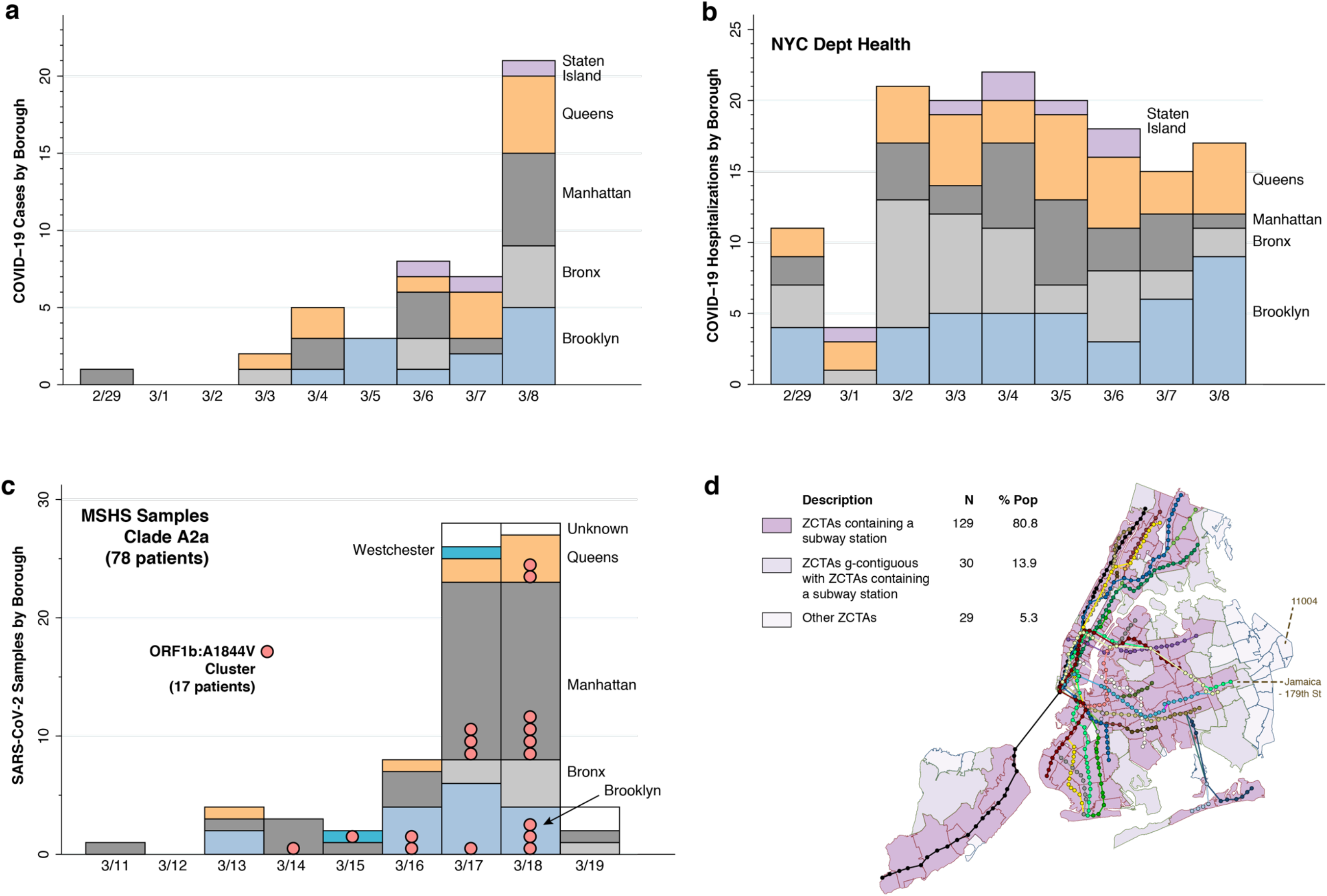
Evidence of Early Rapid, Widespread Community Transmission. **a**. Counts of the earliest cases of test-confirmed COVID-19 reported by the NYC health department, starting on February 29, 2020.^10^ The counts represent individuals initially identified through targeted testing of symptomatic persons in accordance with restricted criteria issued on February 28 by the U.S. Centers for Disease Control (CDC).^28^ The horizontal scale indicates the dates that the cases were diagnosed over the ensuing 8 days. **b**. Timeline of the numbers of individuals ultimately diagnosed with COVID-19 in connection with their inpatient hospitalizations, derived from the same data source.^10^ The counts of these hospitalization are graphed according to each individual’s date of admission during the same 9-day interval. **c**. Timing and locations of 78 viral isolates from dominant clade A2a collected from patients of the Mount Sinai Health System (MSHS) in New York.^20^ In addition to four of the New York City boroughs (Brooklyn, Bronx, Manhattan, and Queens), two of the MSHS A2a patients were from Westchester County (colored cyan) and five patients had unknown residence (colored white). Pink bubbles denote a cluster of 17 samples sharing a common point mutation, A1844V in open reading frame (ORF) 1a. **d**. Map of all subway lines and stops in NYC, distinguishing 129 zip code tabulation areas (ZCTAs) containing a subway station, 30 ZCTAs geographically contiguous with a ZCTA containing a subway station, and 29 other ZCTAs. The Jamaica – 179^th^ Street station at the end of the F Line connects to the 43 bus-route running along Hillside Avenue, which terminates in ZCTA 11004.

**Fig. 2:**
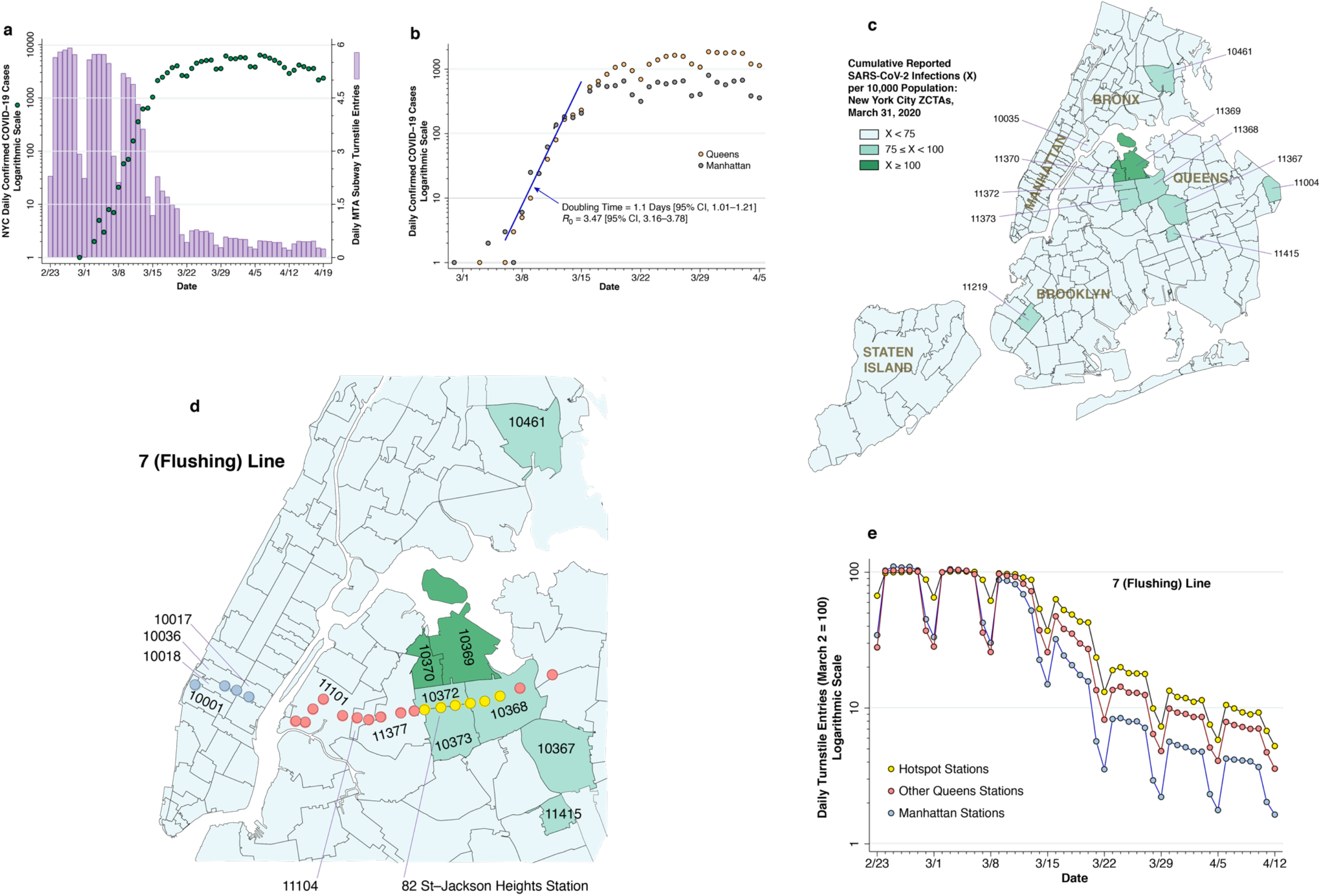
Subway Volume and COVID-19 Cases. **a**. COVID-19 case counts and subway volume during February 23 – April 19, 2020. The dark green-colored data points show the numbers of daily, city-wide confirmed COVID-19 cases reported by the NYC health department,^10^ measured on a logarithmic scale at the left. The lilac-colored bars show the daily volume of trips on the city’s subway system, computed from the Metropolitan Transportation Authority (MTA) turnstile data^22^ and measured on a linear scale at the right. **b**. COVID-19 case counts in Manhattan and Queens during March 1 – April 5, 2020, with a common, initial exponential growth at a doubling time of 1.1 during the week of March 8–15, as estimated by Poisson regression, followed by divergence of the epidemic paths in the two boroughs. **c**. Zip code tabulation areas (ZCTAs) in New York City, color coded according to cumulative case incidence as of March 31, 2020, showing a high-incidence hot spot in the Queens-Elmhurst area. **d**. Section of Fig. 2c, overlaid by the locations of the 22 stations of the 7 (Flushing) subway line, including those in Manhattan (sky blue), the hot spot (yellow), and the remainder of Queens (pink).^38^ The 82^nd^ Street – Jackson Heights station within the yellow group is identified for reference. The pink-colored Mets-Willets station within ZCTA 11368 is on the other side of Grand Central Parkway. **e**. Relative numbers of daily turnstile entries for each of the three zones of the 7 (Flushing) line identified in Fig. 2d. The daily turnstile entries, likewise derived from the MTA turnstile data ^22^, were normalized so that the volume on Monday, March 2 was equal to 100 for each zone. As of March 16, the subway entries into the yellow hotspot stations were 63.2 percent of their March 2 level, while entries into the remaining Queens stations and Manhattan stations were, respectively, 47.7 and 32.2 percent of their March 2 baseline.

**Fig. 3:**
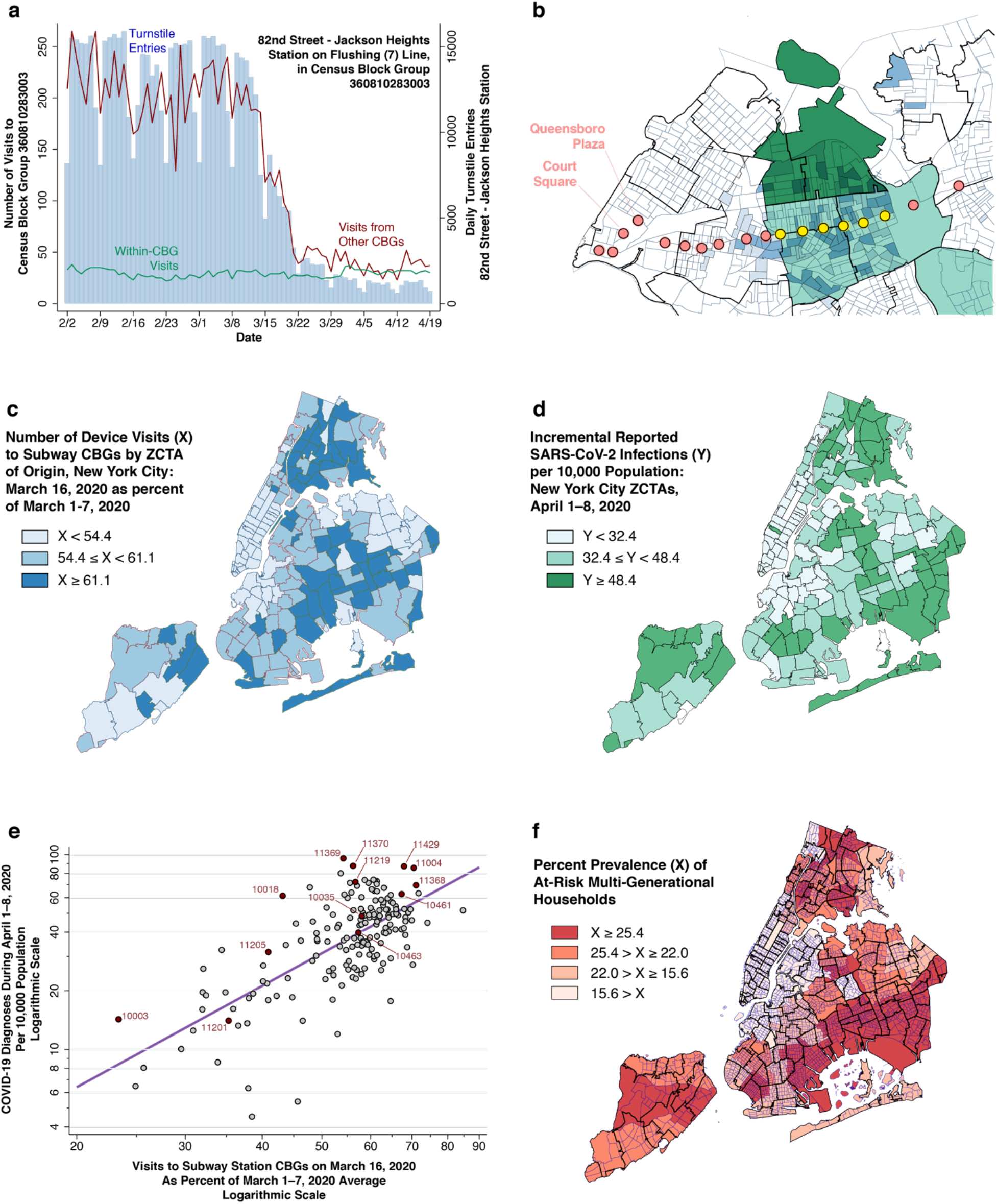
Smartphone Device Movements and COVID-19 Cases. **a**. Daily turnstile entries into the 82^nd^ Street – Jackson Heights Station (blue vertical bars, right axis) and numbers of smartphone device visits to the census block group (CBG) containing that station (red and green lines, left axis). The green data series shows the number of visits from devices originating within the same CBG, while the red data series shows the number of visits from devices originating in other CBGs. **b**. Section of Queens showing CBG boundaries within ZCTA boundaries, overlaid with locations of stations along the 7 (Flushing) subway line. Two-tiered light-dark blue shading identifies those origin CBGs with the highest number of combined visits to a pair of destination CBGs along the 7 (Flushing) line: one of the yellow hotspot stations and one station in the Queensboro Plaza-Court Square commercial complex. In addition, those ZCTAs within the Queens-Elmhurst hot spot have been shaded light-dark green according to the two-tiered color scheme of Figs. 2c and 2d. **c**. Number of device visits to subway CBGs on March 16, expressed as a percent of visits during March 1–7, 2020. **d**. Incidence of newly diagnosed COVID-19 cases during April 1–8, 2020. **e**. Incremental COVID-19 incidence during April 1–8 (vertical axis) related to the number of visits to subway CBGs on March 16, 2020 (horizontal axis). Each point in the log-log plot is an individual ZCTA. As in Fig. 3c, visit counts are normalized so that average volume during the first week of March equaled 100. The superimposed line is the ordinary least squares fit. (See Supplement.) **f**. Prevalence of at-risk multi-generational households, measured as the proportion of households in each ZCTA with at least 4 persons, of whom at least one person was 18–34 years of age and at least one other person was at least 50 years of age.^27^ The map shows census tract boundaries within ZCTA boundaries. Color scheme reflects quartiles of prevalence.

### Population Data

Data on the total populations of zip code tabulation areas (ZCTAs) were derived from the Census Bureau’s American Community Survey 5-year estimates for 2015–2019, accessed from the data server at the Missouri Census Data Center.^11^ Data on the total populations of census block groups (CBGs) were likewise derived from the Census Bureau’s American Community Survey 5-year estimates for 2015–2019, accessed from the Census Bureau’s website.^12^

### Geography

The Metropolitan Transportation Authority (MTA) website for developers ^13^ was our source for the geocoordinates (longitude and latitude) of each of the subway stations, including the 22 stations on the Flushing Local (Number 7) line, as depicted in Figs. 1d, 2d, 3b, and 4b.

**Fig. 4:**
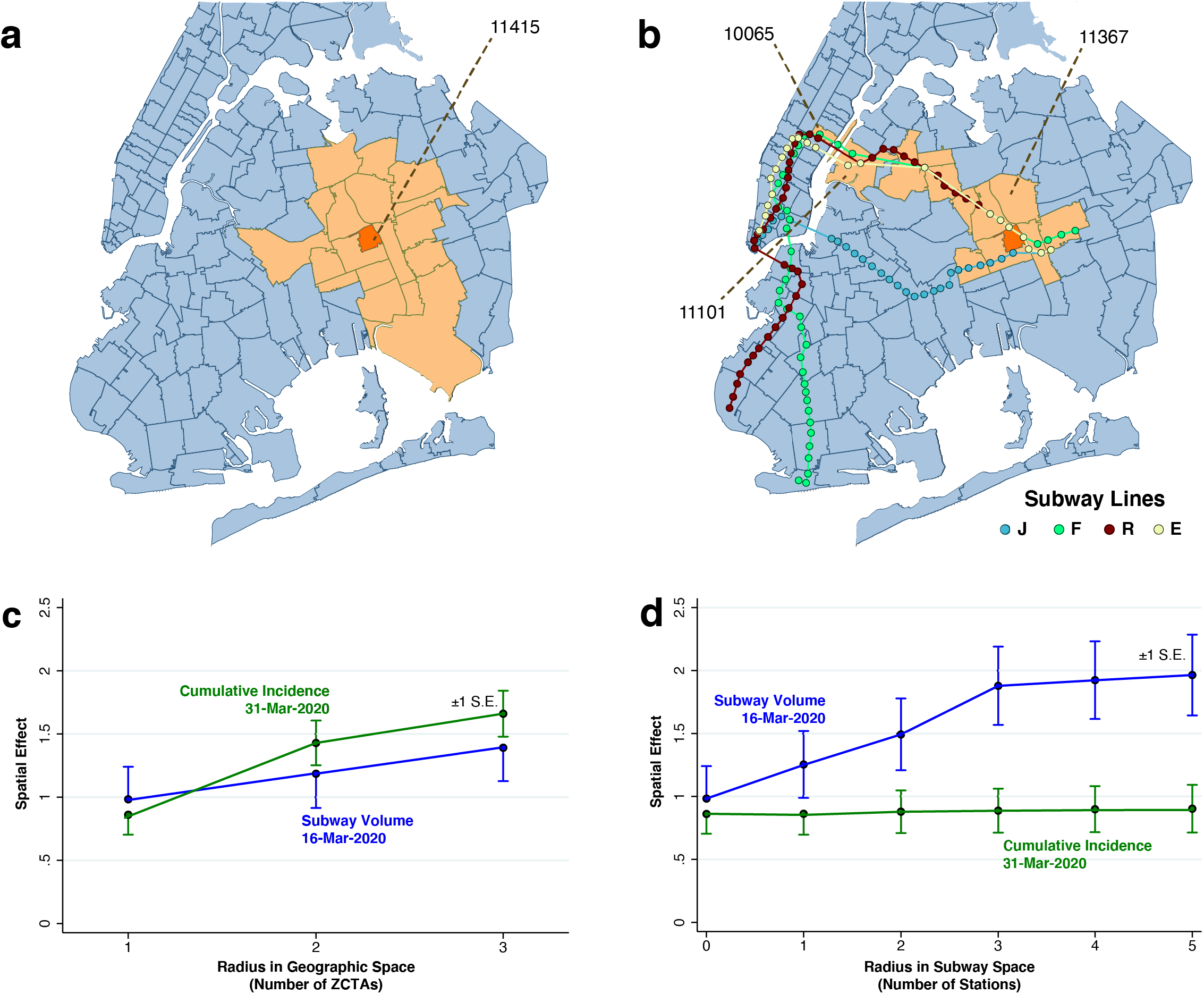
Spatial Analysis. **a**. Map of ZCTA 11415 (colored orange), surrounded by 18 ZCTAs (colored peach) within a geographic radius of 2 ZTCAs. **b**. Map of ZCTA 11415 (colored orange), along with 12 ZCTAs either within a geographic radius of 1 ZCTA or a subway radius of 5 station stops. **c**. Estimated spatial effects of cumulative incidence through March 31 and subway volume on March 16 in relation to radius in geographic space, as the contiguity criterion was varied from g to g+g^2^ to, g+g^2^+g^3^. Cumulative incidence exhibited a significantly increasing trend. (In a 2-sided z-test comparing a radius of 3 with radius of 1, *p* < 0.001.) Subway volume did not. (In an analogous 2-sided z test, *p* = 0.268.) **d**. Estimated spatial effects of cumulative incidence through March 31 and subway volume on March 16 in relation to radius in subway space, as the contiguity criterion varied from g, to g+s, to g+s+s^2^, up to g+s+s^2^+s^3^+s^4^+s^5^. Subway volume exhibited a significantly increasing trend. (In a 2-sided z-test comparing a radius of 3 with radius of 0, *p* = 0.026.) Cumulative incidence did not.

We downloaded the polygon shapes of all census block groups (CBGs) in New York City from the Census Bureau’s website.^14^ We relied on the *Stata* program *geoinpoly*,^15^ which uses a ray-casting algorithm to determine whether a point is contained in a polygon, to identify the unique CBG containing each subway station (as illustrated by the 82^nd^ St–Jackson Heights station in Figs. 2d and 3a).

To map CBGs into ZCTAs, we proceeded in four steps. First, we used *Stata* mapping software to verify that most CBGs were uniquely contained in a given ZCTA (Supplement Fig. A). Second, we employed *QGIS* software to compute the centroids of each CBG in New York City based upon the Census Bureau’s polygon shape files. Third, we downloaded the polygon shape files of all ZCTAs from the New York City health department’s data archive.^16^ Finally, we employed *geoinpoly* once again to determine the ZCTA shape polygon that contained the centroid of each CBG.

As discussed in detail below, our analysis of the prevalence of at-risk multi-generational households relied upon the Census Bureau’s American Community Survey Public Use Microdata Sample (PUMS) for the 5-year period 2015–2019.^17^ The data records for the PUMS are identified at the level of the Public Use Microdata Area (PUMA) ^18^, which is an aggregate of census tracts, which are in turn aggregates of CBGs. To map PUMAs into ZCTAs, we downloaded the scheme for aggregating New York City census tracts into PUMAs from the ESRI’s ArcGIS Hub,^19^ which in turn gave us a mapping from PUMAs to CBGs. We then relied on our prior mapping of CBGs into ZCTAs to go from PUMAs directly to CBGs, as seen in Supplement Fig. A.

### Data on Phylogenetic Analysis of Viral Isolates

To construct Fig. 1c below, we relied upon two data sources: (a) the tab entitled *Clade A2a GISAID IDs* within in the spreadsheet *Data File S2*, posted in the Supplementary Materials of Gonzalez-Reiche et al.;^20^ and (b) the spreadsheet *Supplementary Table 2*, posted in the Supplementary Materials of a later study of COVID-19 patients treated within the New York University Langone Hospital system.^21^ We merged the two files on the unique common identifier variable *gisaid_epi_isi* (where GISAID stands for Global Initiative on Sharing All Influenza Data). This gave a total of 78 MSHS viral samples authored by Gonzalez-Reiche et al. within the A2a clade, including date, location and *strain* identifier. These 78 samples formed the database for the vertical bars in the figure.

Next, we used the variable *strain* in the merged file to identify the 17 virus samples specifically highlighted as sharing the ORF1b:A1844V mutation in the *New York Cluster 1* in Figure 2C of Gonzalez-Reiche et al.^20^ These 17 samples are indicated as the pink bubbles in Fig. 1c. This mutation resulted from a single amino acid substitution from alanine (A) to valine (V) at position #1844 in the stretch of the virus’ RNA coding for its ORF1b protein, which is one of the two replicase proteins common to SARS coronaviruses. In terms of the virus’ underlying genetic code, the mutation corresponded to a single base substitution in the virus’ positive-sense mRNA codon from G<u>U</u>X to G<u>C</u>X, where G = guanine, U = uracil, C = cytosine, A = adenine, and X = any of these four bases. This single RNA base substitution (or missense mutation) was shared by samples of infected persons residing in Manhattan, Queens, Brooklyn and Westchester County, collected during the space of only five days (March 14–18).

### Data on Subway Turnstile Entries

The data on turnstile entries were similarly derived from the MTA’s website for developers.^22^ Since stations typically have multiple turnstiles, and since the turnstile counters are updated at intervals during each day, computation of entries by station and by date involved the aggregation of data points across large data sets with millions of individual observations. Accurate coding required us to take account of the fact that some turnstiles ran backwards, while others were reset when they reached their numerical limit. Still, the city-wide temporal patterns seen in Fig. 2a are consistent with other independent estimates.^23^

### Classification of Flushing Line (Local 7) Stations

In Figs. 2d and 2e below, we classified subway stations along the 7 (Flushing) Line into three groups: The six key stations within the Queens-Elmhurst hot spot, indicated in yellow from west to east, were: 74th St – Broadway; 82nd St – Jackson Hts; 90th St – Elmhurst Av; Junction Blvd; 103rd St – Corona Plaza; and 111 St. The stations within Manhattan, indicated in sky blue from west to east, were: 34th St – Hudson Yards, Times Sq – 42nd St, 5th Ave – Bryant Pk, and Grand Central – 42nd St. The remaining stations within the borough of Queens are indicated in pink.

### Data on Smartphone Device Movements

Our data on smartphone device movements come from the Social Distancing database maintained by SafeGraph.^24^ Every device movement (or *visit*) had an *origin* and a *destination*. Each device’s unique origin was the CBG where it regularly spent the night. Every CBG in which the device stopped for more than 1 minute during a 24-hour period was counted as the destination of a visit, but the duration of each visit was not recorded. The 1-minute cutoff was chosen by SafeGraph; it was not under the researcher’s control. For each calendar day and each CBG of origin, the database recorded the number of devices that visited each destination CBG. A destination CBG can be the same as the origin CBG.

We tested whether smartphone device movements whose destination CBG contained a subway station could serve as a proxy for subway turnstile entries. For each station, we compared two time series: the number of visits to the destination CBG containing that subway station, which we’ll call *subway CBG visits*, and the number of turnstile entries into that station. This comparison is illustrated for a particular subway station in Fig. 3a.

We further investigated the origins of those smartphone devices whose destination CBGs contained one of the six key stations within the Queens-Elmhurst hot spot. For each CBG, we determined two visit counts. The first count, which we denote *n*_1_, accumulated the total number of visits originating in that CBG with a destination at any one of the six stations during the months of January and February 2020. (The 74^th^ Street–Broadway station on the 7 (Flushing) line shared the same CBG as the Jackson Heights–Roosevelt Ave. station on the intersecting 6^th^ Avenue Local (M) line.) The second count, which we denote *n*_2_, accumulated the total number of visits originating in the same CBG during the same interval with a destination at either the Queensboro Plaza or Court Square stops, two of the principal destinations within the Queens portion of the 7 (Flushing) line. We then ranked each origin CBG by the statistic *n*_*min*_ = *min*{*n*_1_, *n*_2_}, which captured trips to and from the Queens-Elmhurst yellow stations and the Queensboro Plaza–Court Square complex. In Fig. 3b, the lighter-shaded CBGs correspond to 100 > *n*_*min*_ ≥ 50, while the darker shaded CBGs correspond to *n*_*min*_ ≥ 100.

To estimate subway visits by ZCTA, we aggregated the number of device visits to all destination CBGs containing a subway station, and then further aggregated these CBG-specific counts of subway visits at the ZCTA level. Supplement Fig. A illustrates the congruence between CBGs and ZCTAs. Supplement Fig. B illustrates the temporal evolution of visits to all subway station CBGs originating from four specific ZCTAs: 10003 (Manhattan), 11201 (Brooklyn), 11205 (Brooklyn), and 11368 (Queens).

Our reconstruction of the origins of subway visits from smartphone mobility data is to be distinguished from prior studies relying instead upon the SafeGraph Patterns Schema, a separate database which classifies visits by their destination points of interest.^25 26^ The latter database did not categorize subway stations as a point of interest.

### Prevalence of At-Risk Multi-Generational Households

We relied upon the five-year (2015–2019) public use microsample of the U.S. Census Bureau’s American Community (ACS)^17^ to estimate the proportion of households in New York City that were at risk for multi-generational transmission of SARS-CoV-2. Following an earlier study of intra-household transmission in Los Angeles County,^27^ we defined an at-risk household as having at least 4 persons, of whom at least one person was 18–34 years of age and at least one other person was at least 50 years of age. Based upon a subsample of 148,686 New York City households in the 5-year ACS database, we found that 18.3 percent of households satisfied this criterion. Across 55 public use microdata areas (PUMAs), the median proportion of at-risk households was 22.0 percent, with the 25^th^ and 75^th^ percentiles at 15.6 and 25.4 percent, respectively. As described above, we then mapped the PUMA-specific estimates into ZCTAs. Across 176 ZCTAs, the median proportion of households at risk was 22.4 percent, with the 25^th^ and 75^th^ percentiles at 13.7 and 24.8 percent, respectively. The minimum proportion was 3.2 percent (ZCTA 10017 in Manhattan), while the maximum proportion was 35.8 percent (11414 and 11420 in Queens).

### Contiguity in Geographic and Subway Space

Our concepts of geographic and subway contiguity, including an accompanying formal matrix algebra, are developed in detail in the Supplement. Briefly, the map of ZCTAs in New York City can be regarded as a finite set of *M* > 0 compact polygons in a two-dimensional plane, indexed by *i* = 1, …, *M*. No two ZCTAs share any interior points in common, but they can share boundary points. When ZCTAs *i* and *j* do share boundary points, we say that they are geographically contiguous, or *g-contiguous*. By contrast, when ZCTA *j* is the next stop after ZCTA *i* on some subway line in some direction, *w*e say that ZCTAs *i* and *j* are contiguous in subway space, or *s-contiguous*. G-contiguity does not imply s-contiguity, nor does s-contiguity imply g-contiguity.

As further elaborated in detail in the Supplement, we formulated compound relationships based on the elemental notions of g- and s-contiguity. To illustrate compound g-contiguity, Fig. 4a shows all ZCTAs that are (g+g^2^)-contiguous with ZCTA 11415. Equivalently, the figure displays all ZCTAs within a geographic contiguity radius of 2. To illustrate compound s-contiguity, Fig. 4b displays all ZCTAs that are (g+s+s^2^+s^3^+s^4^+s^5^)-contiguous with ZCTA 11415, that is all ZCTAs that are either g-contiguous with that ZCTA or within a subway radius of 5 stops along the same or a connecting line. In general, compound g- and s-contiguity accommodate a variable radius.

### Non-Spatial Regressions

Let *y* denote a *M* × 1 column vector of ZCTA-specific observations of incremental COVID-19 incidence during April 1–8, 2020 (mapped in Fig. 3d). Let *X*_0_ denote the corresponding ZCTA-specific column vector of observations on the cumulative incidence of COVID-19 as of March 31 (Fig. 2c). Let *X*_1_ denote the corresponding vector of observations on relative subway volume as of March 16, 2020 (Fig. 3c), and let *X*_2_ denote the prevalence of at-risk multigenerational households (Fig. 3f). As detailed in the Supplement, we estimated non-spatial models of the form log *y* = *α* + *β*_0_ log *X*_0_ + *β*_1_ log *X*_1_ + *β*_2_ log *X*_2_, where the logarithm is assumed to operate separately on each vector coordinate.

### Spatial Regressions

We then considered spatial regression models of the form log *y* = *α* + *β*_0_ log *X*_0_ + *β*_1_ log *X*_1_ + *β*_2_ log *X*_2_ + *γ*_0_ log *WX*_0_ + *γ*_1_ log *WX*_1_ + *γ*_2_ log *WX*_2_, where *W* is an *M* × *M* spatial weighting matrix. Each contiguity criterion necessarily had its own weighting matrix *W*. As detailed in the Supplement, pre-multiplication of each vector *X*_0_, *X*_1_, and *X*_2_ by *W* computed its respective population-weighted mean value among all ZCTAs satisfying the particular contiguity criterion.

## Results

### Early Rapid, Widespread Community Transmission

Assessment of the extent of infection during the earliest days of the NYC outbreak has been hampered by the initial lack of adequate testing materials. Still, Fig. 1a shows that, despite the narrow testing criteria initially imposed on February 28 by the Centers for Disease Control (CDC),^28^ positive tests had been detected in residents of every borough of the city by March 6. Fig. 1b further demonstrates that by March 1, hospitals had already admitted patients residing in every borough. The incubation period between infection and first symptoms of COVID-19 is 5 days on average,^29^ with a range of up to 2 weeks.^30^ Add to that elapsed time an extra 4–10 more days before a symptomatic individual becomes sick enough to be hospitalized.^31^ Accordingly, in all likelihood, SARS-CoV-2 infections were already occurring by mid-February in every one of the five boroughs of a city of over 8 million inhabitants. This pattern of early rapid, widespread dispersion is sharply distinguishable from the gradual radial geographic expansion of COVID-19 cases observed in the earliest days of epidemic in Los Angeles County,^27^ a comparable sized jurisdiction with 10 million inhabitants.

The data in Fig. 1c help to distinguish between two alternative explanations for this pattern of early rapid, widespread dispersion of SARS-CoV-2 infections: parallel, contemporaneous importation from multiple outside sources; and rapid mixing via community transmission. The figure describes the timing and locations of 78 viral isolates belonging to phylogenetic clade A2a that were collected from patients of the Mount Sinai Health System (MSHS) in New York^20^ soon after the CDC liberalized its testing criteria.^28^ Within this dominant clade, the investigators identified a local transmission cluster with a signature mutation in samples drawn from residents of Brooklyn, Manhattan, Queens, and Westchester County over a 5-day period. This observation goes against parallel seeding from distinct sources as the only explanation.

The evidence from Figs. 1a–c alone does not identify the distinct mechanisms underlying such widespread community transmission in so short an interval. Despite a large body of investigation attempting to retrospectively track down super-spreader events,^32^ the only such documented occurrence is an outbreak of COVID-19 among MTA front-line workers.^33 34^ If only by exclusion, we are left with NYC’s unique subway system,^1^ which, in combination with the MTA’s extensive bus routes,^35^ covers virtually every corner of the city (Fig. 1d).

### Subway Volume and COVID-19 Cases

#### The Collapse of Subway Travel and The Flattening of the Epidemic Curve

For the city as a whole, Fig. 2a compares daily subway turnstile entries^22^ to daily numbers of confirmed COVID-19 diagnoses.^10^ Counts of confirmed cases based on voluntary testing of symptomatic individuals are known to have significantly understated actual numbers of SARS-CoV-2 infections.^36 37^ Still, once the CDC liberalized its testing criteria,^28^ one can see the rapid growth in daily confirmed cases, from 21 on March 8 to 1,038 on March 15.

During that same week from March 8–15, subway volume was already declining from its prior average of 5.6 million turnstile entries per weekday. By the end of that week, daily COVID-19 case counts had begun to deviate from their exponential trend. By the time subway rides had fallen to less than one-quarter of their regular volume in the third week of March, the epidemic curve had flattened out. The data are compatible with a causal relation between the drop in subway demand and the deceleration of the epidemic curve, with a delay of 2 ± 1 weeks between the two time-series.

The flattening of the epidemic curve cannot be wholly attributable to official government actions to restrict mobility and reduce interpersonal contact. The decline in subway turnstile entries in Fig. 2a occurred *before* the mayor closed entertainment venues and limited restaurants, bars and cafes to food take-out and delivery on March 17.^39^ While the mayor indeed shut down nightclubs, movie theaters, and concert halls, no one ordered the subways closed. To the contrary, state and local officials attempted to quell the public’s rising fears about the risks of coronavirus transmission on public transit.^40 41^ A more plausible explanation is that voluntary action motivated by fear of contagion – and not a response government coercion – precipitated the collapse of subway demand, which at least in part contributed to the subsequent flattening of curve.

#### The Attenuated Decline in Subway Use and the Emergence of Hotspots

If the decline in subway use in fact caused the observed deceleration of the epidemic in Fig. 2a, then those areas of the city with a more rapid decline would experience a greater deceleration, while those areas with an attenuated decline would experience continued epidemic growth. This prediction is tested in Figs. 2b through 2e, where we focus on an emerging hotspot in the Elmhurst area of Queens and the specific subway line running through it.

Fig. 2b plots daily confirmed COVID-19 cases over time in two boroughs: Manhattan and Queens.^10^ During the week starting March 8, the case counts from both boroughs followed an exponential path with a slope of 0.63/day, which, based on a generation time of 5.5 days,^42^ implies a basic reproductive number ℛ_0_= 0.63 × 5.5 = 3.47. This estimate of ℛ_0_ is comparable to that estimated for the outbreak in Wuhan,^43^ a city with its own massive subway system,^44^ but higher than that estimated for Italy.^45^ By the third week in March, however, the two incidence curves began to diverge significantly. By the last full week of the month, weekday reported cases in Manhattan were down to about 600, while weekday reported cases in Queens exceeded 1,500.

Fig. 2c maps the cumulative incidence of confirmed COVID-19 cases according to zip code tabulation area (ZCTA) as of March 31, 2020. While there are isolated high-incidence ZCTAs in Brooklyn and the Bronx, there is a notable cluster in the Elmhurst area in Queens, especially ZCTAs 11369 and 11370, where the cumulative incidence of confirmed cases had already exceeded 1 percent of the population. Manhattan, by contrast, shows no foci of cumulative COVID-19 incidence in excess of 0.75 percent of the population. Comparison of the borough-level data in Fig. 2b suggests that the Queens-Elmhurst hotspot seen in Fig. 2c may have begun to emerge in the third week of March.

Fig. 2d displays the 22 stations of the 7 (Flushing) subway line^38^ overlaid on a section of the map of Fig. 2c. Fig. 2e shows that turnstile entries into the six yellow-colored stations within the Queens-Elmhurst hotspot remained significantly higher than the remaining pink Queens and blue Manhattan stations, especially from the week of March 15 onward. The divergence in the decline in subway volume among these three groups is consistent with the prediction that the attenuated decline in turnstile volume promoted continued epidemic spread of SARS-CoV-2.

### Smartphone Device Movements and COVID-19 Incidence

#### Smartphone Device Movements as a Proxy for Subway Turnstile Entries

The turnstile volume data in Figs. 2a and 2e show how many riders entered the subway system at various stations, but not where these subway riders originated. To fill this data gap, we relied on data on the movements of smartphones equipped with location-tracking software.^24^

Each smartphone movement (or “visit”) had a recorded origin and a destination census block group (CBG). Fig. 3a illustrates how we reproduced the daily pattern of turnstile entries into each subway station by adding up those smartphone visits whose *destination* was the CBG where that station was located. That finding allowed us to rely upon smartphone visits to station CBGs as a proxy for turnstile entries, and thus to study the *origins* of subway visitors.

Fig. 3b illustrates how smartphones entering the 7 (Flushing) Line at the yellow stations (already identified in Fig. 2d) originated not only from the ZCTA where the station was located, but also from the adjacent high-incidence ZCTAs. This finding suggested that we could reliably estimate the number of subway visits originating from each ZCTA by adding up subway-station smartphone visits that originated from that ZCTA. To that end, Fig. 3c displays the resulting map of the estimated distribution of subway visits by ZCTA as of March 16, 2020, expressed as a percentage of the corresponding baseline volume during the first week of March.

### Relation Between Subway Visits in Mid-March and COVID-19 Incidence in Early April

Fig. 3d maps the incidence of newly diagnosed COVID-19 cases per 10,000 during April 1–8, 2020. If smartphone visits are in fact a reliable proxy for subway visits, and if an attenuated decline in subway visits in certain areas of the city in mid-March resulted in the subsequent emergence of high-incidence hotspots in those areas by early April, then we should expect to observe a strong correlation between the visit volume mapped in Fig. 3c and the incidence mapped in Fig. 3d. This prediction is in fact borne out in the bivariate plot of Fig. 3e. The slope of the ordinarily least squares-fitted line was significant at the level *p* < 0.001.

The significant bivariate association in Fig. 3e held up in multivariate models that took account of two additional ZCTA-specific covariates: (i) cumulative COVID-19 incidence through March 31 (already mapped in Fig. 2c); and (ii) the prevalence of multi-generational households (mapped in Fig. 3f), a well-established ecological determinant of transmission rates.^27 46-48^. In all such multivariate models, the estimated parameters were significantly different from zero at the level *p* = 0.006 or lower. (See Supplement Table A.)

### Spatial Analysis

#### Geographic and Subway Contiguity

The foregoing multivariate models of COVID-19 incidence during the first week of April do not account for the possibility of contagion across ZCTAs. While models of the spatial propagation of SARS-CoV-2 across geographic units have been proposed and tested,^27 49^ New York City presents a potentially unique example of contagion in *subway space*, as opposed to geographic space.

To that end, consider two distinct ZCTAs, abstractly labeled *i* and *j*. We say that ZCTAs *i* and *j* are geographically contiguous, or simply *g-contiguous*, when they share at least one common boundary point. By contrast, the same two ZCTAs are subway contiguous, or simply *s-contiguous*, when ZCTA *j* is the next stop after ZCTA *i* on some subway line in some direction. As detailed in the section “Contiguity in Geographic and Subway Space” in the Supplement, these elemental relations between ZCTAs can be compounded. For example, two ZCTAs *i* and *j* are *g*^*2*^*-contiguous* if there is a third distinct ZCTA labeled *k*, such that ZCTA *i* is g-contiguous with ZCTA *k* and ZCTA *k* is in turn is g-contiguous with ZCTA *j*.

As a further extension of the concept of compound contiguity, we say that two ZCTAs are (*g+g*^*2*^*)-contiguous* if they are *either* g-contiguous *or* g^2^-contiguous. This situation is illustrated in Fig. 4a, which shows ZCTA 11415 (colored orange) in Queens, surrounded by a total of 18 ZCTAs (colored peach) that are (g+g^2^)-contiguous with ZCTA 11415. Within this group, four ZCTAs are g-contiguous with ZCTA 11145, while the remaining 14 ZCTAs are situated effectively within a radius of 2 from the reference ZCTA 11415.

Fig. 4b, by contrast, displays 12 ZCTAs (again colored peach) that are (g+s+s^2^+s^3^+s^4^+s^5^)-contiguous with the reference ZCTA 11415 (again colored orange). ZCTA 11367 is exclusively g-contiguous with the reference ZCTA 1145. The remaining ZCTAs are accessible within 5 subway stops along the same or a connecting line. Thus, ZCTA 11101 in Queens is accessible via four stops on the E Line, while ZCTA 10065 in Manhattan is further accessible after a transfer at the Queens Plaza station to the R, N or W Lines.

#### Spatial Regressions: Subways, Networks, and Percolation

Our non-spatial regression models permitted us to measure how prior conditions in a particular ZCTA (including March 16 subway volume and March 31 cumulative cases) influenced subsequent COVID-19 incidence during April 1–8 within the *same* ZCTA. By contrast, our spatial regression models (detailed in the Supplement) permitted us to measure how COVID-19 incidence during the first week of April was influenced by prior conditions in *other* ZCTAs. To implement these spatial models, we did not arbitrarily allow each ZCTA to be influenced by all other ZCTAs, but instead restricted the radius of potential contagion in both geographic and subway space. Thus, for a particular ZCTA in Queens, Fig. 4a illustrates a limited *geographic* radius of 2 ZCTAs, while Fig. 4b illustrates a limited *subway* radius of 5 stops.

We repeatedly estimated such between-ZCTA spatial effects as we varied the allowable radius of influence – from 1 to 3 in geographic space, and from 0 to 5 in subway space. As we enlarged the allowable radius of influence in geographic space, as shown in Fig. 4c, we found that cumulative incidence in other ZCTAs as of March 31 became an increasingly strong predictor of subsequent COVID-19 incidence, whereas the volume of subway visits originating in other ZCTAs as of March 16 did not. On the other hand, as we enlarged the allowable radius in subway space, as shown in Fig. 4d, we found just the reverse. That is, the volume of subway visits originating in other ZCTAs on March 16 was an increasingly strong predictor of subsequent COVID-19 incidence during the first week in April. Cumulative incidence in other ZCTAs as of March 31, by contrast, showed no such trend in relation to the allowable radius in subway space.

Our finding that subway volume as of March 16 exhibited increasingly contagious effects in subway space supports the conclusion that SARS-CoV-2 was being propagated via a subway-based network at least through March 16. Our finding that March 31 cumulative incidence exhibited increasingly contagious effects in geographic space supports the conclusion that percolation of new cases through local geographic spread had subsequently become the dominant mode of propagation by the end of March. Once local clusters developed, further percolation of new cases via transmission within multi-generational households (Fig. 3f) became dominant.

## Discussion

The evidence presented here supports three distinct but not mutually exclusive hypotheses. First, the subway system played a critical role in the rapid, widespread community transmission of SARS-CoV-2 infection throughout New York City during late February and early March 2020. Second, the ensuing marked decline in subway travel was an important mechanism by which the public’s growing perception of risk was translated into reduced community transmission of the virus. Third, those areas with an attenuated decline in subway use subsequently became hotspots of viral infection in late March and early April 2020.

One alternative interpretation is that subway travel was no more than a proxy for other determinants of vulnerability to COVID-19. In higher-risk communities, so the argument goes, many residents had service jobs that could not be performed remotely. Such an interpretation, however, does not square with the spatial-effect findings in Fig. 4d, which imply some mechanism of contagion running along subway lines. A more responsive counterargument would have to assign at least an indirect role to the subway system. Thus, the decline in turnstile entries seen in Figs. 2a and 2e (and Supplement Fig. B) could have reflected employees’ responses to their employers’ requests to work from home, which in turn reduced workplace exposure, where contagion would in fact have taken place. This version does not require that infected individuals transmitted their infections inside subway cars or on station platforms. It concedes only that public transport was an efficient vehicle for moving infected individuals from the periphery of the city to its commercial centers and back again many times a day.

This last counterargument, however, does not square with the evidence on the known mechanisms of SARS-CoV-2 transmission. An infected person exhales moist air containing very small droplets loaded with the virus.^50^ A passenger without a mask standing two feet away from an infected rider without a mask for just 15 minutes would almost certainly have inhaled virus particles, even if the infected rider never coughed or sneezed.^51^ An infected person constantly sheds virus particles in the form of fomites on almost every surface he touches, such as glasses, keys and phones.^52^ That would include the stainless-steel poles shared by standing passengers. Social distancing can be difficult if not impossible in crowded subway cars and platforms, as well as in public transportation conveyances and transportation hubs generally.^53^ A crowded subway train or platform would thus have been an ideal incubator for coronavirus transmission. In a study of outbreaks involving three or more cases in municipalities in China outside Hubei Province, transport-based transmission was second only to home-based transmission.^54^ The extensive outbreak among MTA front-line workers (and later, their family members) has no alternative explanation.^33 34^

Yet another counterargument draws upon conflicting studies of the transmission of other respiratory viruses in public transport. A study of the London Underground offered supporting evidence of the transmission of influenza-like illness.^55 56^ But a simulation study calibrated to the 1957–1958 flu epidemic in New York City estimated only a small contribution from subway travel.^57^ A cross-sectional study of 121 cities found a negative association between public transit use and mortality from pneumonia and influenza during 2006-2015.^58^ In contrast to a basic reproductive number of ℛ_0_= 3.47 (95% confidence interval, 3.16–3.78) for SARS-CoV-2 in New York City estimated here, seasonal influenza has an ℛ_0_ in the range of 1.2–1.4, while pandemic influenza has an ℛ_0_ in the range of 1.4–1.8, with the high end representing the 1918 pandemic.^59^ While a wave of COVID-19 cases swept through the U.S. during October 2020– January 2021, reported diagnoses of influenza A and B were way down.^60^ The relevance of studies of influenza in public transport is, at the very least, questionable.

The evidence presented here also highlights the methodological limitations of alternative approaches to studying the role of the subways in the propagation of SARS-CoV-2. The test conducted in Figs. 2b–2e demonstrates the importance of studying *changes* in subway volume during the course of the COVID-19 outbreak. Less informative would be a study relating COVID-19 rates to static survey data on the proportion of individuals in each ZCTA regularly riding public transit prior to the epidemic. Our results also point to the importance of conducting tests of causation when baseline subway volume and COVID-19 incidence are high. A finding that coronavirus cases no longer relate to subway volume once subway use has plummeted to below 10 percent of baseline reveals little if anything about what happened back in March. The map of the Flushing Local line in Fig. 2b further highlights the pitfalls of studies that assign the entire volume of turnstile entries into a subway station to its enclosing ZCTA.^7 8^ Such a procedure, which effectively assumes that only people who live in the same ZCTA take the local subway, would erroneously discard the high-incidence ZCTAs 11369 and 11370, which have no subway within their boundaries.

If we are to successfully control future pandemic threats – and, for that matter, future outbreaks of COVID-19 – we need to understand in exhaustive detail how SARS-CoV-2 first took hold and then established hot spots in major urban epicenters throughout the world. Considerable effort has been made to understand exactly what happened in Wuhan.^43 61^ A study of Los Angeles County has tracked the initial seeding of imported infections in affluent areas as it spread radially to high-density neighborhoods, where the virus percolated through multi-generational households.^27^ While the outbreak in Italy has been traced phylogenetically to the Lombardy region, it remains unclear how exactly it started out and spread.^62^ A more recent phylogenetic study of viral samples from New York state during March–May 2020 confirmed the importance of Queens as a major transmission hub and provided supporting evidence of widespread geographic dispersion.^63^ Only 22 percent of the samples from New York City, however, were collected before the last week of March.^64^ Numerous investigators have relied upon compartmental models to understand the early dynamics of SARS-CoV-2 outbreaks.^26 27 43 65^ The evidence presented here for New York City points instead to a model of network-wide transmission followed by local percolation of infections.^66-69^

If the subway system indeed played a critical role in the early propagation of SARS-CoV-2, as supported by the evidence assembled here, we need to understand that the conventional methods of personal contact tracing are less likely to be useful in halting future outbreaks. That means more sophisticated contact tracing through the pings of mobile devices and records of electronic transactions will be necessary.^70 71^ To that end, the MTA will need to adopt a new system of digital passes, already in use in many cities worldwide, which would permit investigators to find out more than just the crude number of turnstile-clicks at each station.

In advance of the next outbreak, we will need to know whether the subways served principally as a rapid spatial disseminator of externally acquired infections,^5 6 72^ or as significant locus of in situ transmission.^4^ In the former case, social distancing and mandatory face coverings would not alone stop the rapid, widespread seeding of infections throughout the five boroughs that we observed in February and March of 2020. In the latter case, we will need to study now whether a policy of running only express lines with limited density might be a feasible alternative to the complete cordon sanitaire adopted in Wuhan more than a year ago.^73^

## Data Availability

This study relies on several large databases that are publicly accessible via the Internet. The Reference section contains the URLs for access to the underlying data on COVID-19 cases and hospitalizations, subway turnstile entries, and phylogenetic analyses of virus samples. The underlying data on smartphone movements, provided by SafeGraph, are available to researchers only by permission. We have posted our data analyses at the Open Science Framework (OSF) in a project entitled New York City COVID-19 Epidemic.

https://osf.io/v7k23/

## Acknowledgments

This article represents the sole opinion of its author and does not necessarily represent the opinions of the Massachusetts Institute of Technology, Eisner Health, or any other organization or individual.

## Competing Interests Declaration

The author has no competing interests to declare.

## Funding Declaration

The author has no funding sources to declare.

## Human Subjects Declaration

This study relies exclusively on anonymized, publicly available data that contain no individual identifiers.

## Data Availability Statement

This study relies on several large databases that are publicly accessible via the Internet. The Reference section contains the URLs for access to the underlying data on COVID-19 cases and hospitalizations, subway turnstile entries, and phylogenetic analyses of virus samples. The underlying data on smartphone movements, provided by SafeGraph, are available to researchers only by permission. We have posted our data analyses at the Open Science Framework (OSF) in a project entitled *New York City COVID-19 Epidemic* (https://osf.io/v7k23/).

## Notes

### Competing Interest Statement

The authors have declared no competing interest.

### Summary of Updates

Figure 4 revised. Additional reference #72 added. Methods section moved up before Results. Clarifying changes in Methods and Results sections. Supplemental file remains unchanged.

