## Supplement for "Critical Role of the Subways in the Initial Spread of SARS-CoV-2 in New York City"

Jeffrey E. Harris MD PhD* July 3, 2021

**Additional Details of the Construction of Subway Visit Volume by ZCTA of Origin**

#### Overlay of Census Block Groups on Zip Code Tabulation Areas

Fig. A overlays several maps of a section of New York City comprising parts of Manhattan, Queens, and Brooklyn. The bottom layer, indicated by the blue subdivisions, shows the boundaries of individual census block groups (CBGs). The middle layer, indicated by the black subdivisions, shows the boundaries of zip code tabulation areas (ZCTAs). The top layer, indicated by the colored regions, identifies the Queens-Elmhurst hotspot ZCTAs shown in main text Figs. 2c and 2d. Most CBGs were fully contained within specific ZCTAs. In those cases where a CBG overlapped more than one ZCTA, we assigned the CBG to the ZCTA that contained the centroid of the CBG.

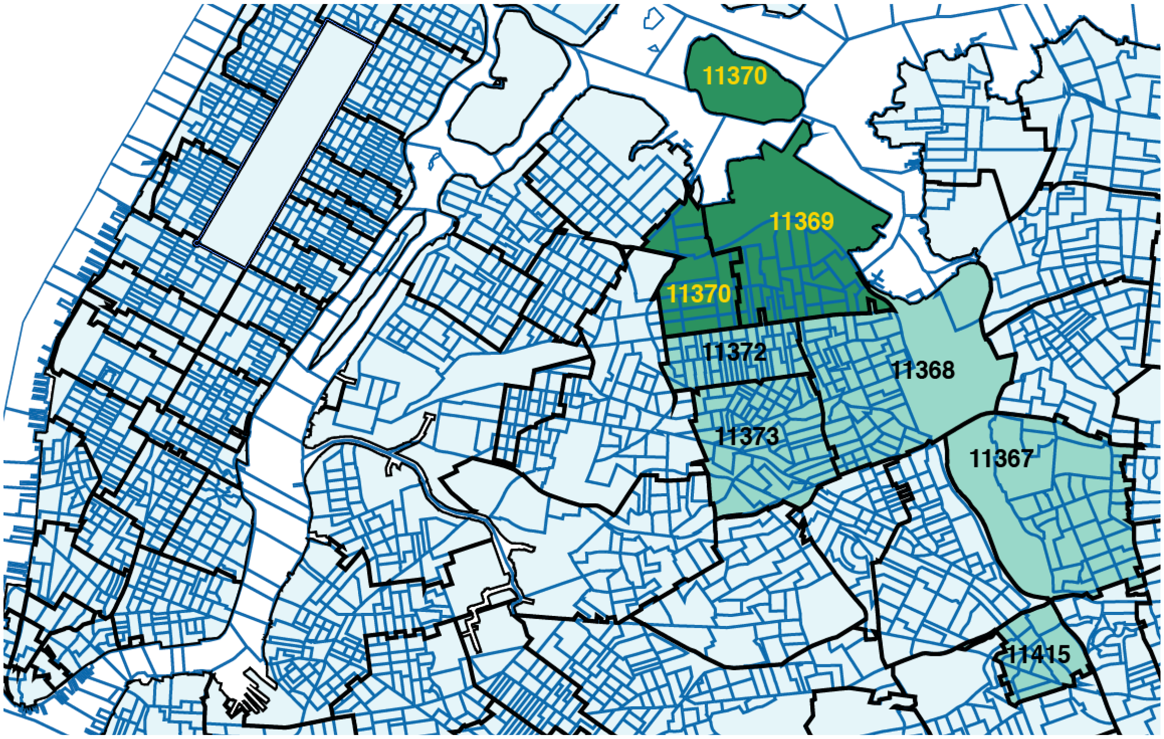


*Fig. A. Overlay of ZCTAs on CBGs in a section of Manhattan and Queens.*

#### Temporal Evolution of Visits to Subway Station CBGs Originating from Four ZCTAs

Fig. B shows the temporal evolution of visits to subway station CBGs originating from four specific ZCTAs: 10003 (Manhattan), 11201 (Brooklyn), 11205 (Brooklyn), and 11368 (Queens). The volume of visits to subway station CBGs has been expressed as a percentage of the volume for the first week in March. The dispersion in subway visit volume can been seen along the vertical dashed line located at March 16. The specific percentages at that date are: 23.38 for 10003; 35.28 for 11201; 40.92 for 11205; and 71.08 for 11368.

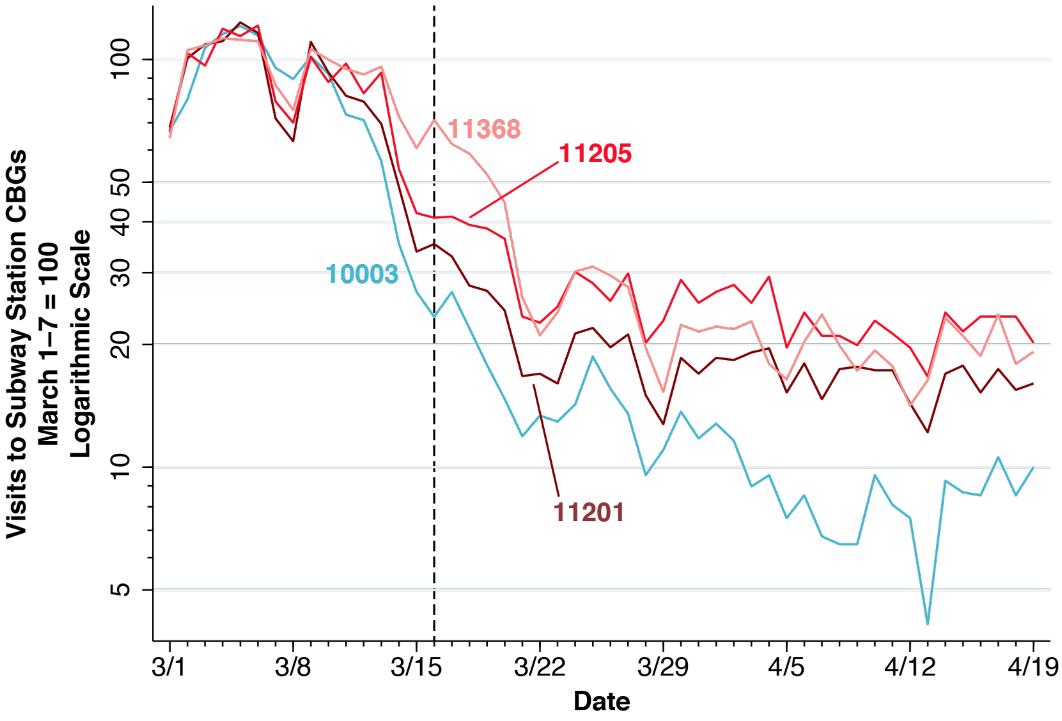


*Fig. B. Temporal evolution of visits to subway station CBGs originating from four ZCTAs.*

### Contiguity in Geographic and Subway Space

#### G-Contiguity

We think of our map of ZCTAs in New York City as a finite set of $M > 0$ compact polygons in a two-dimensional plane, indexed by $i = 1,\ldots,M$ . No two ZCTAs share any interior points in common, but they can share boundary points. When ZCTAs $i$ and $j$ do share boundary points, we say that they are geographically contiguous, or simply *g-contiguous*. By convention, we don't allow a ZCTA to be g-contiguous with itself, so that g-contiguity, considered as a binary relation, is symmetric, but it is neither reflexive nor transitive. To take a simple example, Fig. C shows three geographic areas, that is, $M = 3$. Area #1 is g-contiguous with area #2, but neither #1 nor #2 is g-contiguous with #3.

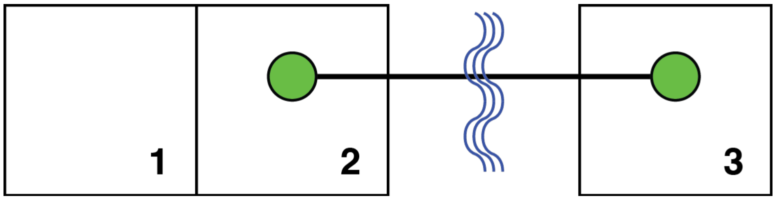


*Fig. C. Three geographic areas. Areas #2 and #3 are connected by a bidirectional subway line, which crosses a body of water.*

Geographic contiguity can be represented by an $M \times M$ symmetric square matrix $G$ with binary elements, where element $g_{ij} = 1$ if and only if area $i$ is g-contiguous with area $j$, and where $g_{ij} = 0$ otherwise. Since we do not allow an area to be g-contiguous with itself, the diagonal elements of $G$ are all zeros. For the example of Fig. C, the g-contiguity matrix takes the form $G = \begin{bmatrix} 0 & 1 & 0 \\ 1 & 0 & 0 \\ 0 & 0 & 0 \end{bmatrix}$, where the rows and columns are ordered in accordance with labeling in the figure.

#### S-Contiguity

We say that two areas $i$ and $j$ are contiguous in subway space, or simply *s-contiguous*, when area $j$ is the next stop after area $i$ on some subway line in some direction. Once again, we don't allow an area to be s-contiguous with itself. So long as subway trains run in both directions, s-contiguity is likewise a symmetric relation, though it is not reflexive or transitive. In Fig. C above, area #2 is s-contiguous with area #3, but these two areas are not g-contiguous.

Fig. C helps us understand why g- and s-contiguity are distinct relations. The figure specifically captures the case where two areas are physically separated by a body of water. This is the case for a number of ZCTAs in Manhattan, which are connected by subway lines to stops in Brooklyn, Queens, and Staten Island. The other important case where g- and s-contiguity are divergent is where a subway line has express trains that skip over many local stops.

Subway contiguity can be analogously represented by a symmetric square matrix *S*, where element $s_{ij} = 1$ if and only if area *i* is s-contiguous with area *j*, and $s_{ij} = 0$ otherwise. By convention, the diagonal elements $s_{ii}$ are all zero. For the example of Fig. C, the s-contiguity matrix takes the form $S = \begin{bmatrix} 0 & 0 & 0 \\ 0 & 0 & 1 \\ 0 & 1 & 0 \end{bmatrix}$.

#### Compound Geographic Continuity

In Fig. D below, we have removed the body of water separating areas #1 and #2 in Fig. 1, and have added area #4, which is both g- and s-contiguous with area #3. The respective g- and s-contiguity matrices now take the form $G = \begin{bmatrix} 0 & 1 & 0 & 0 \\ 1 & 0 & 1 & 0 \\ 0 & 1 & 0 & 1 \\ 0 & 0 & 1 & 0 \end{bmatrix}$ and $S = \begin{bmatrix} 0 & 0 & 0 & 0 \\ 0 & 0 & 1 & 0 \\ 0 & 1 & 0 & 1 \\ 0 & 0 & 1 & 0 \end{bmatrix}$.

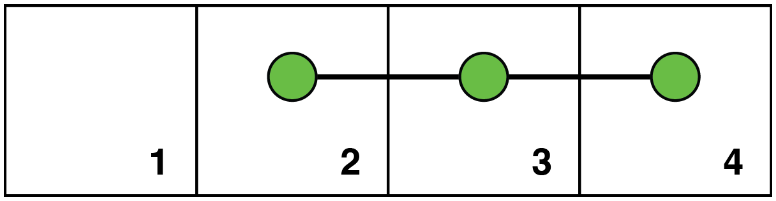


*Fig. D. Four geographic areas. Areas #2, #3, and #4 are connected by a bidirectional subway line. The body of water has been removed, so that area #1 is g-contiguous with area #2.*

Fig. D allows us to consider compound forms of contiguity. In particular, we say that two areas are *g²-contiguous* if one can travel from the first to the second in two steps, going through a third area that is a common geographic neighbor with both. In Fig. D, we can see that area #1 is $g^2$-contiguous with area #3, and area #2 is $g^2$-contiguous with area #4.

Intuition suggests that the corresponding $g^2$-contiguity matrix would be $G^2 = GG$, that is, the matrix product of *G* with itself, but that is not exactly the case. For the configuration in Fig. D, we would have $G^2 = \begin{bmatrix} 1 & 0 & 1 & 0 \\ 0 & 2 & 0 & 1 \\ 1 & 0 & 2 & 0 \\ 0 & 1 & 0 & 1 \end{bmatrix}$. The problem is that the diagonal elements of $G^2$ count the number of ways one can travel in two steps back to the area where one started.

To form the corresponding contiguity matrix, we first need to define a mapping $b:\Omega \to \Omega$ from the space $\Omega$ of $M \times M$ matrices onto itself with the following properties. For any matrix $Z \in \Omega$ with typical element $z_{ij}$, we have: (i) $b(z_{ii}) = 0$; and (ii) for all $i \neq j$, $b(z_{ij}) = \text{sign}(z_{ij})$, where $\text{sign}(z)$ equals 1 if $z$ is strictly positive and 0 otherwise. Then the matrix representing the g²-contiguity relation in Fig. D is $b(G^2) = \begin{bmatrix} 0 & 0 & 1 & 0 \\ 0 & 0 & 0 & 1 \\ 1 & 0 & 0 & 0 \\ 0 & 1 & 0 & 0 \end{bmatrix}$.

We can use our matrix algebra to define other compound forms based upon the elemental relation of g-contiguity. For example, an element on row *i* and column *j* of the matrix $b(G + G^2)$ is equal to 1 if and only if area *i* is *either* g-contiguous *or* g²-contiguous with area *j*. In that case, we would get the matrix $b(G + G^2) = \begin{bmatrix} 0 & 1 & 1 & 0 \\ 1 & 0 & 1 & 1 \\ 1 & 1 & 0 & 1 \\ 0 & 1 & 1 & 0 \end{bmatrix}$.

#### Compound Subway Continuity with Multiple Subway Lines

It would seem natural to define s²-contiguity in a manner entirely analogous to g²-contiguity. That is, area *i* would be s²-contiguous with area *j* if there is a distinct area *k* such that area *i* is s-contiguous with area *k*, and area *k* is s-contiguous with area *j*. But that approach runs into an ambiguity when there are multiple, disconnected subway lines. In Fig. E below, the green line continues to run from area #2 to area #3, just as in Fig. D, but a separate red line now runs from area #3 to area #4. Under our original definition, area #2 is s-contiguous with area #3, and area #3 is s-contiguous with area #4, and so area #2 would be s²-contiguous with area #4.

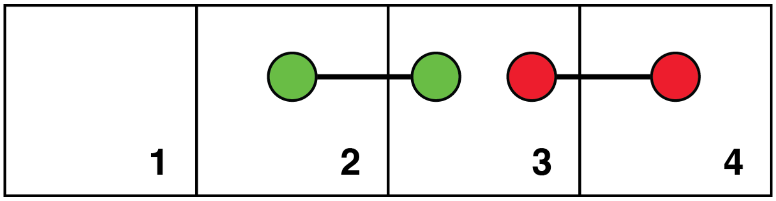


*Fig. E. Four geographic areas. Areas #2 and #3 are connected by the bidirectional green line, while areas #3 and #4 are separately connected by the bidirectional red line.*

To address this issue, we say that area *i* is *$s^2$-contiguous* with area *j* if there exists a distinct area *k* such that area *k* is the next stop after *i* on some subway line and area *j* is the next stop after area *k* along t*he same or a connecting* subway line. We use the term connecting to refer to cases where a rider can change to another line by transferring within the same station. Under this clarified definition, areas #2 and #4 are $s^2$-contiguous in Fig. D, but the same two areas are not $s^2$-contiguous in Fig. E.

To formalize these notions, let *T* be an $N \times N$ square matrix, where $N > 0$ is the number of distinct, non-connected subway stations. The typical element $t_{ij}$ equals 1 if stations *i* and *j* are successive stops along the same or a connecting subway line. Otherwise, $t_{ij} = 0$. Let *R* be an $N \times M$ matrix where typical element $r_{ij}$ equals 1 if subway station *i* is located in area *j*, and $r_{ij} = 0$ otherwise. Then the $M \times M$ matrix representing $s^2$-contiguity is $b\left(R'T^2R\right)$.

Fig. F below illustrates this formalism. In contrast to Fig. 4, we now have $N = 5$ subway stations: A, B and C on the green line, and D and E on the non-connecting red line. The corresponding matrix *T* showing the connected stations has the form $T = \begin{bmatrix} 0 & 1 & 0 & 0 & 0 \\ 1 & 0 & 1 & 0 & 0 \\ 0 & 1 & 0 & 0 & 0 \\ 0 & 0 & 0 & 0 & 1 \\ 0 & 0 & 0 & 1 & 0 \end{bmatrix}$,

where we've ordered the stations alphabetically. The $5 \times 4$ matrix *R* showing which stations belong to which areas assumes the form $R = \begin{bmatrix} 1 & 0 & 0 & 0 \\ 0 & 1 & 0 & 0 \\ 0 & 0 & 1 & 0 \\ 0 & 0 & 1 & 0 \\ 0 & 0 & 0 & 1 \end{bmatrix}$. We thus have

$b\left(R'T^2R\right) = \begin{bmatrix} 0 & 0 & 1 & 0 \\ 0 & 0 & 0 & 0 \\ 1 & 0 & 0 & 0 \\ 0 & 0 & 0 & 0 \end{bmatrix}$. That is, only areas #1 and #3 are $s^2$-contiguous.

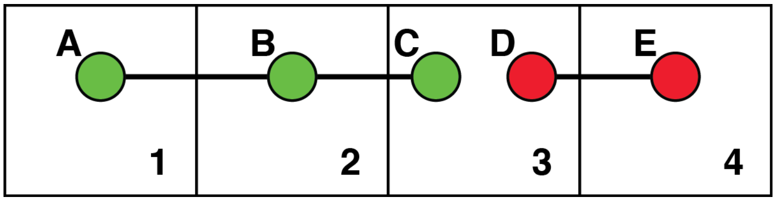


*Fig. F. Four geographic areas. Areas #1. #2 and #3 are connected by the bidirectional green line, while areas #3 and #4 are separately connected by the bidirectional red line.*

As in the case of g-contiguity, we can form other compound relations between areas. Thus, the element in row *i* and column *j* of the matrix $b\left(S+S^2+S^3\right)=b\left(R'\left(T+T^2+T^3\right)R\right)$ equals 1 if area *j* can be reached from area *i* in one, two or three subway stops on the same or connected lines. This compound contiguity matrix is also symmetric.

#### GS-Contiguity

The distribution of blue-shaded census block groups in Fig. 3b in the main text demonstrates that individuals will travel from their ZCTA of origin to a g-contiguous ZCTA to enter a subway station, and then take the subway to work at another ZCTA. Motivated by this finding, we say that area *i* is *gs-contiguous* with area *j* when there exists a third, distinct area *k* such that area *i* is g-contiguous with area *k* and area *k* in turn is s-contiguous with area *j*.

The binary relation defined by gs-contiguity is not symmetric. Thus, in Fig. C above, area #1 is gs-contiguous with area #3, inasmuch a resident of area #1 could travel to the subway station in area #2 and then take the subway one stop over to area #3. But none of the three areas can be gs-contiguous with area #1 because area #1 does not have its own subway stop. Put differently, the matrix $b\left(GS\right)$ has binary elements and zeros along the main diagonal, but it is not symmetric.

Still, we can formulate symmetric contiguity relations combining the two elemental relations. For example, taking advantage of the fact that $SG=\left(GS\right)'$, we can construct the symmetric contiguity matrix $b\left(GS+SG\right)$. The element in row *i* and column *j* of this matrix $b\left(GS+SG\right)$ equals 1 if the two areas are either gs-contiguous, or sg-contiguous. To take a more complex case, the element in row *i* and column *j* of the matrix $b\left(GZ\right)$, where $Z=R'\left(T+T^2+T^3\right)R$, equals 1 if there is another area *k* such that area *i* is g-contiguous with area *k* and area *k* in turn is no more than three subway stops from area *j*. The symmetric matrix $b\left(GZ+ZG\right)$ combines the two relations in both directions.

**Non-Spatial Regressions**

Let $y$ denote an $M\times 1$ column vector of ZCTA-specific observations of incremental COVID-19 incidence during April 1–8, 2020. Let $X_1$ denote the corresponding ZCTA-specific column vector of observations on relative subway volume as of March 16, 2020. The fitted line in Fig. 3e in the main text represents the ordinary least squares (OLS) estimate of the bivariate non-spatial model $\log y=\alpha+\beta_1\log X_1$, where the logarithm is assumed to operate separately on each coordinate of the vectors $y$ and $X_1$. The OLS estimates of the parameters for that model are shown in the row labeled Model 1 in Table A below.

Table A. Non-spatial regressions relating incremental COVID-19 incidence during April 1–8, 2020 ($y$) to the cumulative incidence of COVID-19 through March 31, 2020 ($X_0$), relative subway volume as of March 16, 2020 ($X_1$), and the prevalence of at-risk multigenerational households ($X_2$).*

| Model | $\alpha$ | $\beta_0$ | $\beta_1$ | $\beta_2$ | $R^2$ |
|---|---|---|---|---|---|
| 1 | –3.318<br>( 0.545) | | 1.727<br>(0.135) | | 0.480 |
| 2 | –4.664<br>( 0.460) | 0.757<br>(0.079) | 1.346<br>(0.116) | | 0.664 |
| 3 | –0.321<br>( 0.024) | | 0.631<br>(0.190) | 0.489<br>(0.066) | 0.607 |
| 4 | –1.938<br>( 0.517) | 0.694<br>(0.068) | 0.423<br>(0.151) | 0.426<br>(0.053) | 0.756 |

* All models entailed $N$ = 176 observations. Standard errors are shown in parentheses below each parameter estimate. The estimate of $\alpha$ in Model 3 was not significantly different from 0. Otherwise, all parameter estimates were significantly different from zero at the level $p$ = 0.006 or lower.

We now let $X_0$ denote the corresponding ZCTA-specific column vector of observations on the cumulative incidence of confirmed COVID-19 cases as of March 31, 2020, mapped in Fig. 2c. We further let $X_2$ denote the corresponding ZCTA-specific column vector of observations on the prevalence of at-risk multigenerational households, mapped in Fig. 3f. Then we can also estimate multivariable non-spatial models of the form $\log y = \alpha + \sum_{v=0}^{2} \beta_v \log X_v$,

where some of the parameters $\{\beta_v\}$ can be restricted to equal zero, or all can remain unconstrained. The OLS estimates of the parameters of these alternative non-spatial models are shown in the additional rows labeled Model 2 through 4 in Table A above. In every model, independent of any zero restrictions on the parameters, the estimates of $\{\beta_v\}$ were all significantly different from zero.

Construction of the independent variable $X_1$ relied on the volume of subway CBG visits on March 16, as noted in Fig. B and Fig. 3e in the main text. Varying this cutoff date between March 13 and March 17 did not materially affect our estimates.

**Spatial Regressions**

In addition to these basic cross-sectional, non-spatial models, we considered spatial regression models[1] of the form $\log y = \alpha + \sum_{v=0}^{2} \beta_v \log X_v + \sum_{v=0}^{2} \gamma_v \log WX_v$, where $W$ is an $M \times M$ spatial weighting matrix that may premultiply each of the vectors $X_0$, $X_1$ and $X_2$. This approach accounts for the potential influence of the values of these exogenous variables in geographically or subway contiguous ZCTAs. For example, if we focused on $(g+g^2+g^3)$-contiguity, then coordinate $i$ of the corresponding vector $WX_1$ would measure the population-weighted average subway volume among all ZCTAs within a geographic radius of 3 ZCTAs surrounding ZCTA $i$. We note that we have intentionally avoided the use of a spatially weighted dependent variable $Wy$. Such an approach would amount essentially to correlating contemporaneous values of $y_i$ and $y_j$, an exercise that runs into knotty problems of endogeneity and identification.[1 2]

The spatial weighting matrix $W$ is a population-weighted transform of the corresponding contiguity matrix. Let $u$ denote an $M \times 1$ column vector with coordinate $u_i$ equal to the population of ZCTA $i$, as derived from the Census Bureau's American Community Survey 5-year estimates for 2015–2019,[3] and let $U = \text{diag}(u)$ denote the $M \times M$ diagonal matrix formed from the vector $u$. Consider the elemental case of geographic contiguity, represented by the contiguity matrix $G$. We compute $n = Gu$, where coordinate $n_i$ of the vector $n$ measures the total population of all ZCTAs that are g-contiguous with ZCTA *i*, and let $N = \text{diag}(n)$ denote the $M \times M$ diagonal matrix formed from the vector *n*. Then the corresponding spatial weighting matrix is $W = N^{-1}GU$ . The same procedure would apply to any contiguity matrix other than *G*.

Table B shows the results of a series of three models, where the radius of geographic contiguity was incrementally increased from 1 to 3. In each regression, we excluded three ZCTAs: 10044 (corresponding to Roosevelt Island); 11109 (a very small ZCTA in Queens for which we had no data on smartphone origins); and 99999 (a miscellaneous category that included Central Park, airports, and other non-residential areas). The results are show for the models $\log y = \alpha + \beta_v \log X_v + \gamma_v \log WX_v$, where $v = 0,1$. For both sets of covariates – cumulative incidence through March 31 ( $X_0$ ) and subway volume as of March 16 ( $X_1$ ) – the estimated spatial effect parameter $\gamma_v$ increased as the contiguity radius was expanded. The estimated spatial effect parameters are graphed in Fig. 4d of the main text.

Table B. Spatial Regression Results: Increasing Geographic Radius*

| | Model | | |
|---|---|---|---|
| Parameter | $g$ | $g + g^2$ | $g + g^2 + g^3$ |
| $\beta_0$ | 0.742<br>(0.108) | 0.741<br>(0.093) | 0.793<br>(0.086) |
| $\gamma_0$ | 0.861<br>(0.157) | 1.429<br>(0.177) | 1.661<br>(0.182) |
| $\beta_1$ | 1.110<br>(0.211) | 1.066<br>(0.201) | 0.957<br>(0.195) |
| $\gamma_1$ | 0.983<br>(0.257) | 1.185<br>(0.271) | 1.391<br>(0.264) |

* Standard are errors shown in parentheses below each parameter estimate. All estimates were significantly different from zero at the level $p < 0.001$. All sample sizes $N = 175$. Three ZCTAs were excluded: 10044; 11109; and 99999.

Table C below shows the comparable results of a series of six models, where the radius of spatial contiguity was incrementally increased. To ensure comparability with the results in Table B, all models included the alternative of g-contiguity. An analysis restricted solely to powers of *S* would have resulted in null values of $WX_0$ and $WX_1$ for those ZCTAs that did not contain subway stations. In contrast to Table B, we see a steep gradient only for the spatial effect parameter $\gamma_1$ corresponding to subway volume on March 16, but not for the spatial effect parameter $\gamma_0$ corresponding to cumulative incidence through March 31, 2020. The estimated spatial effect parameters are graphed in Fig. 4c of the main text.

Table C. Spatial Regression Results: Increasing Spatial Radius*

| | Model | | | | | |
|---|---|---|---|---|---|---|
| Parameter | $g$ | $g + s$ | $g + s + s^2$ | IV† | V† | VI† |
| $\beta_0$ | 0.742 (0.108) | 0.747 (0.110) | 0.762 (0.108) | 0.776 (0.108) | 0.812 (0.105) | 0.825 (0.105) |
| $\gamma_0$ | 0.861 (0.157) | 0.863 (0.166) | 0.878 (0.169) | 0.887 (0.175) | 0.899 (0.183) | 0.903 (0.190) |
| $\beta_1$ | 1.110 (0.211) | 0.931 (0.214) | 0.822 (0.216) | 0.676 (0.215) | 0.782 (0.196) | 0.893 (0.185) |
| $\gamma_1$ | 0.983 (0.257) | 1.254 (0.266) | 1.493 (0.285) | 1.880 (0.311) | 1.924 (0.308) | 1.964 (0.321) |

† IV = $g + s + s^2 + s^3$. V = $g + s + s^2 + s^3 + s^4$. VI = $g + s + s^2 + s^3 + s^4$
* Standard are errors shown in parentheses below each parameter estimate. All estimates were significantly different from zero at the level $p < 0.001$ with the exception of $\beta_1$ in Model $G + S^3$ ($p = 0.002$). All sample sizes $N = 175$. Three ZCTAs were excluded: 10044; 11109; and 99999.

We obtained similar results when we included $X_2$ as a covariate in our models (that is, $\log y = \alpha + \beta_v \log X_v + \gamma_v \log WX_v + \beta_2 \log X_2$, where $v = 0,1$). We also obtained qualitatively similar results when we jointly regressed $X_0$ and $X_1$ within the same spatial model (that is, $\log y = \alpha + \beta_0 \log X_0 + \gamma_0 \log WX_0 + \beta_1 \log X_1 + \gamma_1 \log WX_1$).

Finally, we considered a more generalized sequence than that specified in Table C and main text Fig. 4c. This more generalized sequence allows one to transit first to a g-contiguous ZCTA and then take the subway up to a specified number of stops, or in reverse order. More concretely, relying on our matrix algebra above, we define $Z = R'TR$, $Z_2 = R'\left(T + T^2\right)R$, $Z_3 = R'\left(T + T^2 + T^3\right)R$, … We then considered the sequence of contiguity matrices specified by the rule $G$, $b\left(G + Z + GZ + ZG\right)$, $b\left(G + Z_2 + GZ_2 + Z_2G\right)$, $b\left(G + Z_3 + GZ_3 + Z_3G\right)$, … The resulting relation between the radius of subway space and the estimated spatial effect parameters $\gamma_0$ and $\gamma_1$ is shown in Fig. G. The same differential response of the two covariates – cumulative incidence on March 31 ( $X_0$ ) and subway volume on March 16 ( $X_1$ ) – is once again noted.

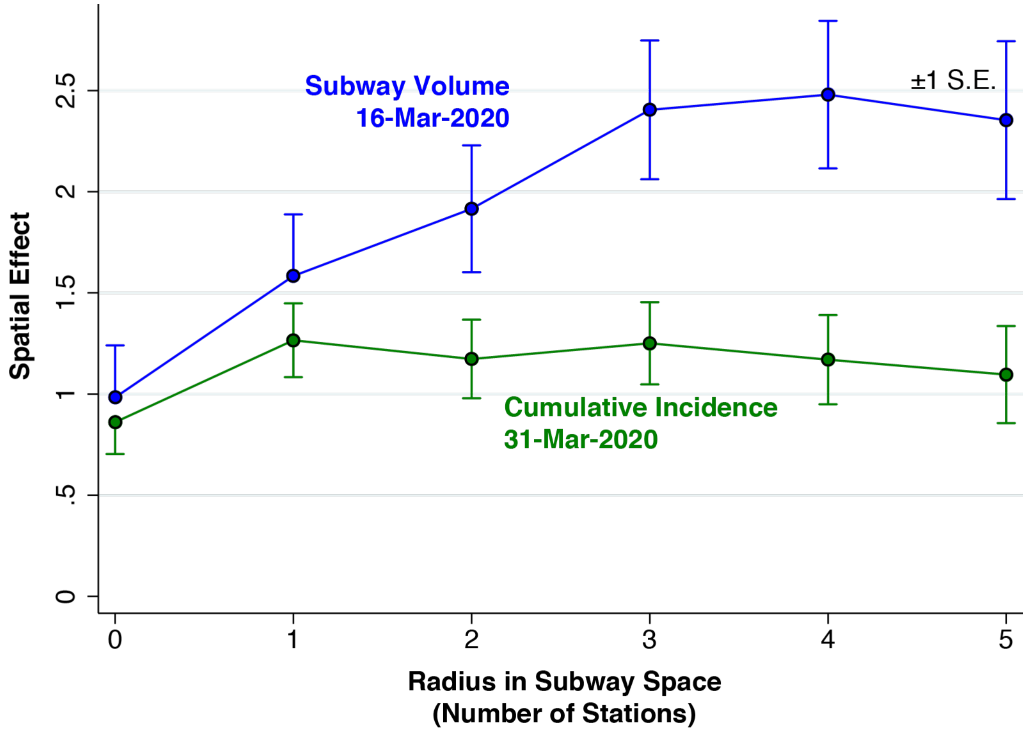


*Fig. G. Relation between radius in subway space and estimated spatial effect parameters in a generalized version of the model of Table C and main text Fig. 4c.*
